# Utility of Point-of-Care Ultrasound in A Resource-Limited Health Care Setting in Southeast Ghana: A Needs Assessment

**DOI:** 10.1101/2023.07.18.23292835

**Authors:** Mahmoud Mohamed Elfadil, Gwen Baraniecki-Zwil, Mobolaji H. Fowose, Julia Schiff, Maham Munawar, Nova Panebianco, Jeffrey A. Kramer

## Abstract

**Background:** Many proven benefits of point-of-care ultrasound (POCUS) are increasingly recognized, including being non-invasive, cost-effective, and of significant diagnostic value. Evidence for these benefits has been replicated in variable settings. However, litle is known about the utilization of POCUS in the developing world and in an out-of-hospital, resource-limited setting. This needs assessment study describes utilization of POCUS in rural Ghana.

**Methods:** We performed retrospective analysis of data from medical missions to Southeast Ghana, evaluating the utilization of POCUS in a rural resource-limited health setting. The first mission trip took place in the Oti region between January 25^th^, 2023, and February 1^st^, 2023. The second was in the Volta region between February 19^th^, 2023, and February 24^th^, 2023. These missions established out-of-hospital remote clinics for walk in patients who are informed about the clinic by the word of mouth by their community leaders and neighbors. We included all POCUS scans performed as part of those missions.

**Findings:** A total of 128 POCUS studies were performed and included in this analysis. Studies were performed for 111 patients (median age 23 years; 49.9% male). Twelve distinct types of sonographic studies were performed. Notably, 48.4% revealed significant findings that helped to confirm a working diagnosis and subsequently formulate or optimize plan of management. A therapeutic intervention was made based on US findings in 16.4% of encounters, while a referral to higher level of care was recommended in 27.3% of cases. Reassurance with ruling out a main differential diagnosis was the case in 7% of encounters.

**Conclusion:** POCUS is effective and can be utilized for a wide range of types of scans effectively in out-of-hospital resource-limited settings in rural Ghana with significant impact on clinical decision-making process.

## BACKGROUND

Ultrasound has been utilized in medical practice since the 1940s^1^. Over the last 8 decades, the technology of medical sonography has evolved rapidly^1^. In recent years, hand-held ultrasound technology has become more popular and affordable^2^. The introduction of smaller portable devices has contributed to increasing utilization of the point-of-care ultrasound (POCUS) in various clinical settings^3^ including global health, especially in resource limited settings^4,5^. The limited availability of diagnostic imaging modalities imposes a challenge for health care systems in low and middle-income countries (LMICs) equally affecting urban and rural areas^5^. Due to lack of medical imaging facilities in LMICs, specific skillsets and competencies has been suggested for healthcare providers to be able to deliver care in those settings^6^, including being trained in POCUS^7^. Notably, practicing POCUS and training medical trainees on it in LMICs was shown to be feasible and effective^7^.

Many proven benefits of POCUS are increasingly recognized, including being non-invasive, cost-effective, and of significant diagnostic value. Evidence for these benefits has been replicated in variable settings for both imaging and guiding medical procedures in primary care as well as critical care and neonatal critical care^8,9^.

In the developing world, main referral centers are often resource-limited and lack the capabilities for medical imaging at a scale that is required to meet the needs of the large catchment areas served. In remote rural areas, the situation is often grimmer. This needs assessment study describes utilization of POCUS in out-of-hospital, resource-limited, remote, rural settings in Ghana. These clinics are often the only opportunity for some to receive medical care. We describe the utility of POCUS in this setting.

## METHODS AND MATERIALS

### Conceptualization and Design

We conducted a retrospective cohort study evaluating the utilization of POCUS in rural resource-limited health settings in Southeast Ghana during two global health, short-term medical missions. The first took place in the Oti region between January 25^th^, 2023, and February 1^st^, 2023. while the second was in the Volta region between February 19^th^, 2023, and February 24^th^, 2023. We analyzed data from POCUS requests and interpretation forms. Data collected included age, gender, indication for the study, findings, diagnosis, and outcomes. The primary objective of this study was to provide a needs assessment of POCUS in the rural, resource-limited healthcare settings in the Oti and Volta regions, Ghana. A secondary objective was to assess the clinical impact of POCUS in these settings.

This study was approved by the University of Pennsylvania Institutional Review Board.

### Population and Settings

The medical mission was initiated by International Needs (Hudsonville, MI) in collaboration with International Needs Ghana (Accra, Ghana) and targeted two regions in the Southeastern Ghana, the Oti and Volta. Remote clinics were hosted in public schools. If required, privacy cubicles were erected for clinical examination and sonographic studies **(Figure 1)**. Patients from all ages were treated at the clinic. Patients were ambulatory and presented from the villages in which they were held. They were informed about the clinics by the word of mouth from their community leaders and neighbors.

**Figure 1.**
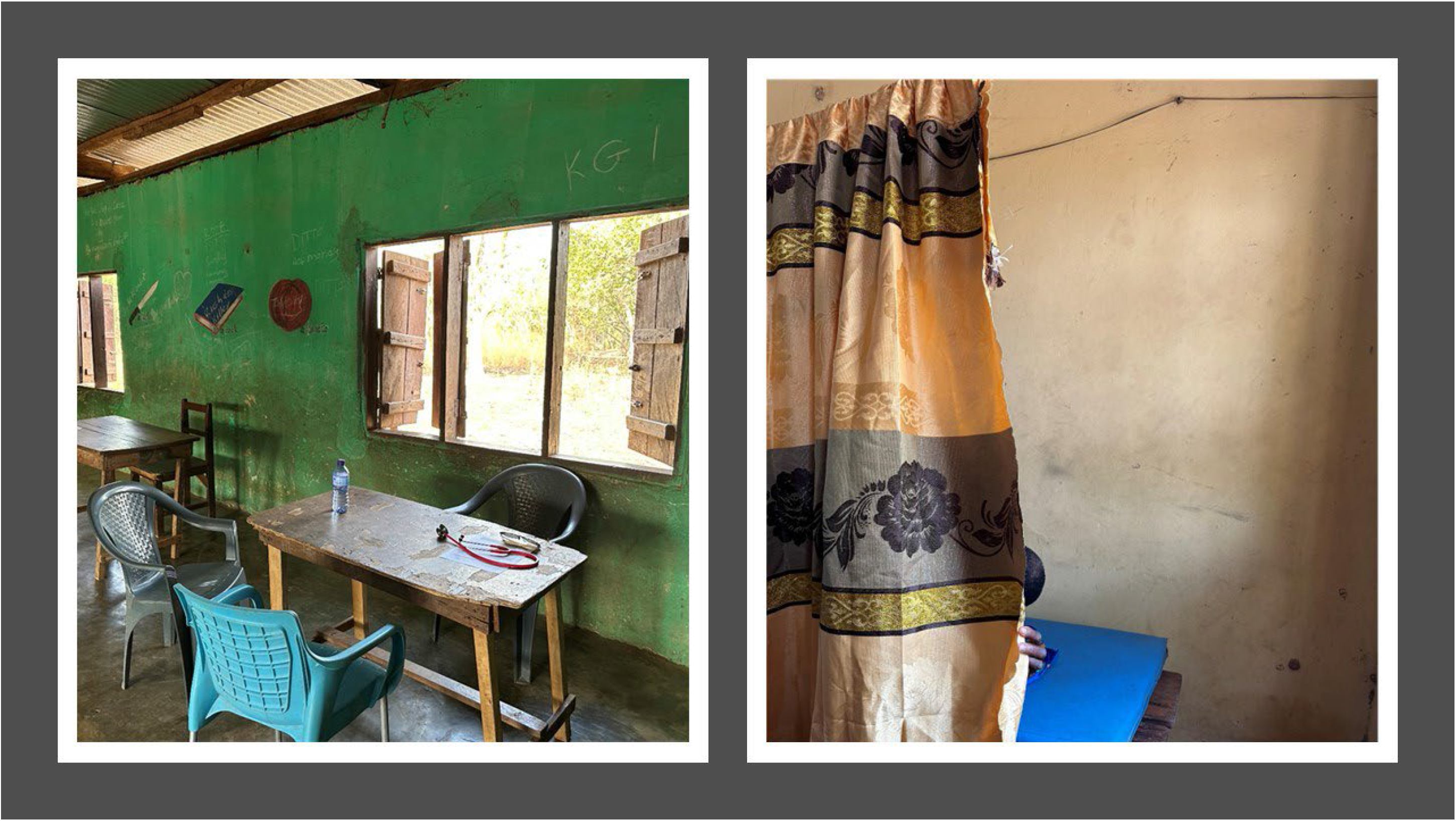
Remote Clinic set-up at the Oti Region (Hosted by a public school).

Scans were ordered by providers physicians participating in the missions. Forms were utilized to request a POCUS study specifying a scan type and indication. The form included some basic demographic information. However, verbal discussions and orders were also used depending on the setting (**Figure 2**). Scans were ordered for various clinical indications.

**Figure 2.**
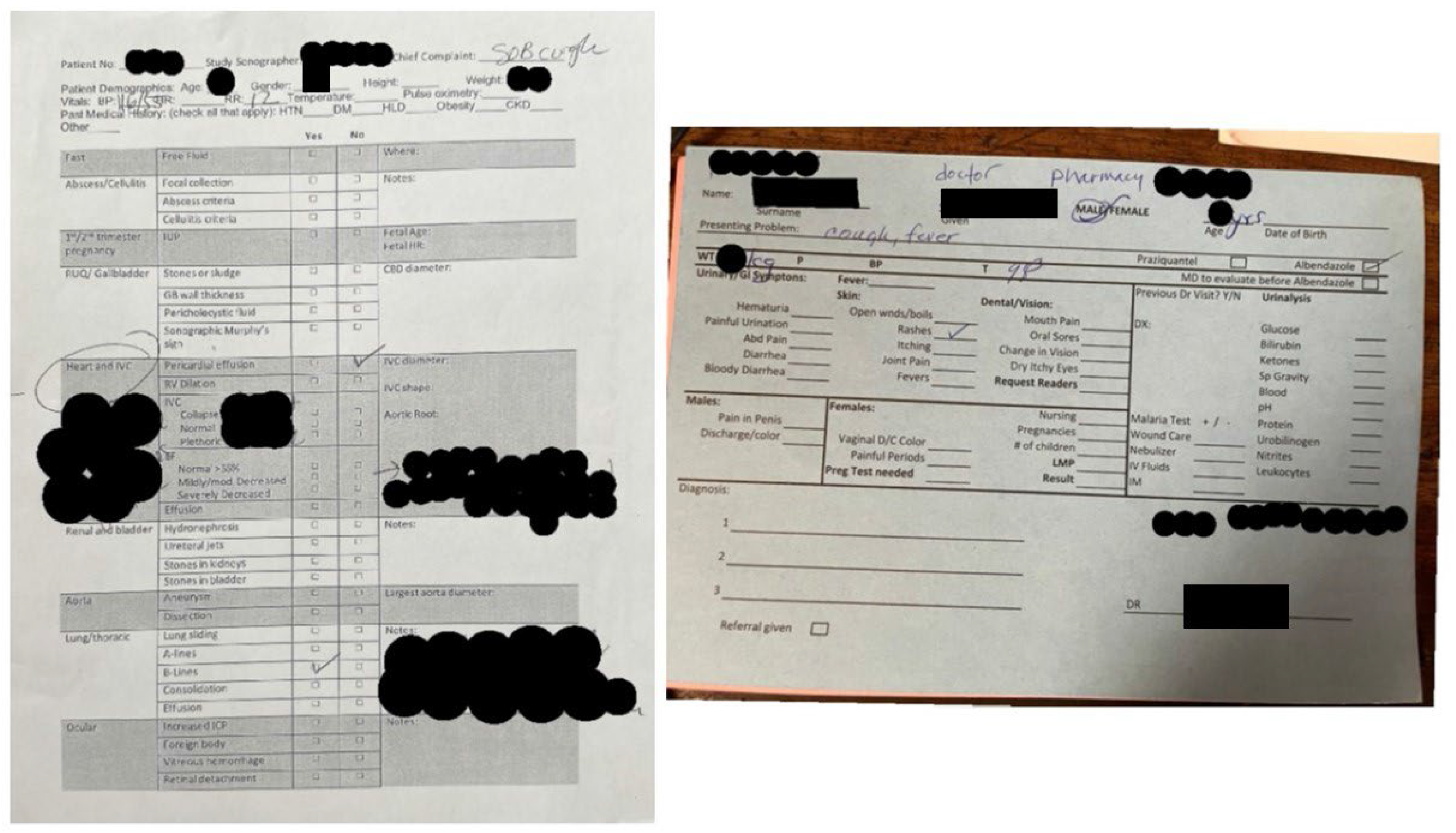
Forms used to request and/or report POCUS studies.

Providers participated in performing POCUS for studies included in this analysis are of variable training and experience background. One provider was a seasoned POCUS trained emergency medicine specialist and POCUS fellowship director, while the other providers included an internal medicine specialist completing POCUS fellowship, an emergency medicine specialist completing global health fellowship, and two emergency medicine residents. These providers also participated in treating a large number of patients in addition to the scans they performed.

In general, our clinics received 4724 patients over the mission period. This number is broken into 2460 patients seen in the Oti region clinics averaging 492 per day, and 2264 patients seen in the Volta region clinics averaging 377 per day.

### Device

All studies were performed using Buterfly iQ® and Buterfly iQ+® (Burlington, MA, USA) hand-held ultrasound device along with smart phone devices.

### Statistical analysis

Descriptive analysis including distribution was performed. Data are presented in frequency and percentages. No contingency analyses were performed given the nature and the scope of the study. Age is presented in median and range as a non-parametric variable.

## RESULTS

A total of 128 POCUS studies were performed and included in this analysis. Studies were performed for 111 patients with 13 patients having more than one study. Patients have median age of 23 (1—80) years and (49.9%) male (**Table 1**). Notably, (35%) of patients were from pediatric age group (age < 18 years). Around two thirds of studies were performed in the Oti region (61.7%), while a third of them (38.3%) were performed in the Volta region in Southeast Ghana (**Figure 3**). Five providers acquired and interpreted all studies independently. Regarding the primary specialty of providers performing the studies, (59.4%) of studies were performed by the Internal Medicine provider, while (40.6%) were performed by an Emergency Medicine provider (**Table 2**).

**Figure 3.**
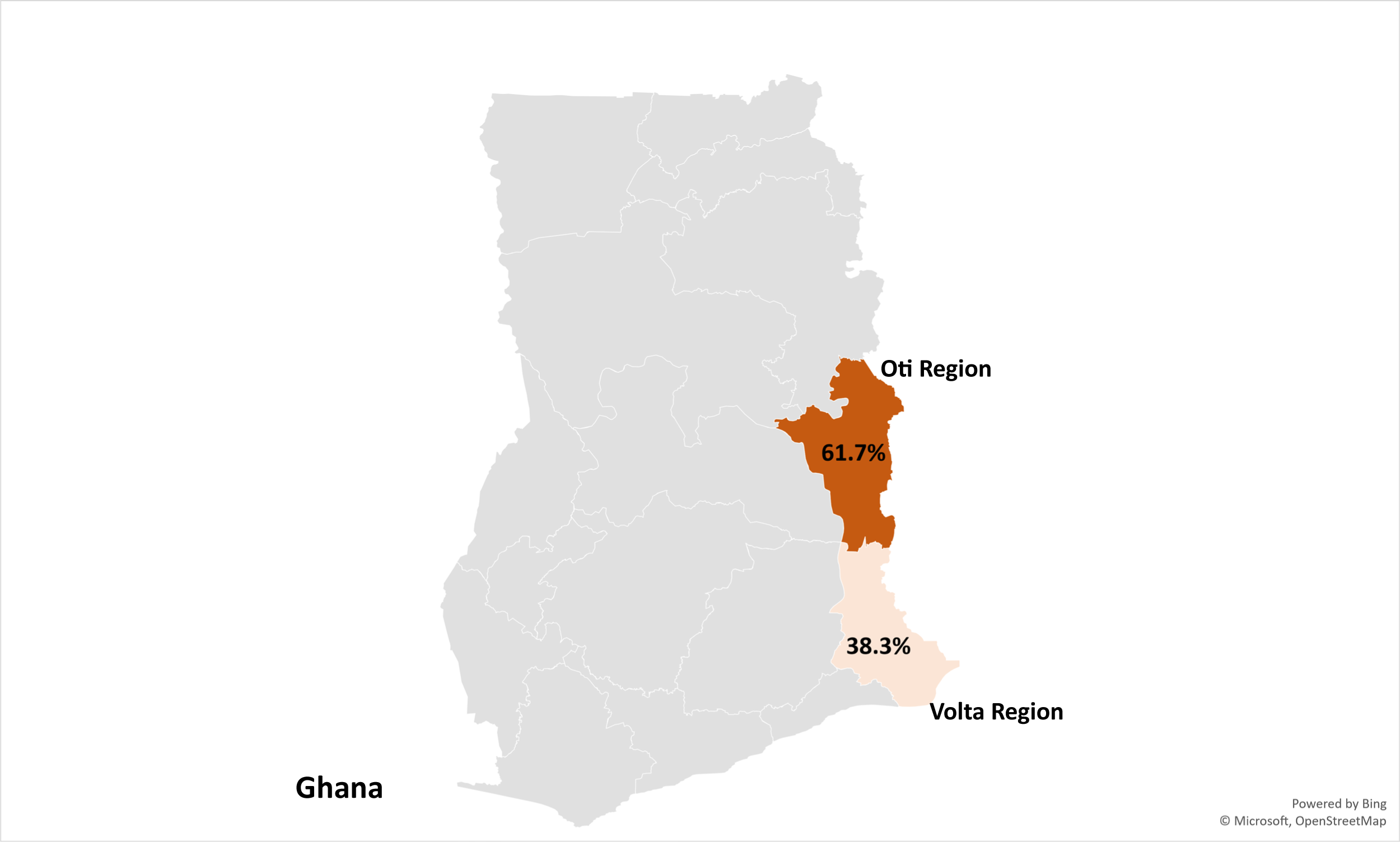
Geographical Distribution of POCUS Studies

**Table 1.** Baseline Demographic and Clinical Characteristics.

| **Table 1. Baseline Demographic and Clinical Characteristics** |  |
| --- | --- |
| **Variables** | **n=111** |
| **Age, year, median (IQR)** | 23 (7—36) |
| **Gender, n (%)** |  |
| ▪ Male | 54 (49.1) |
| ▪ Female | 48 (43.6) |
| ▪ Not reported | 9 (7.3) |
|  | **n=128** |
| **Type of Scan, n (%)** |  |
| ▪ Lung | 36 (28.2) |
| ▪ Renal | 15 (11.7) |
| ▪ Cardiac | 14 (10.9) |
| ▪ 1<sup>st</sup> Trimester Pregnancy | 4 (3.1) |
| ▪ 2<sup>nd</sup> and 3<sup>rd</sup> Trimester Pregnancy | 13 (10.2) |
| ▪ Skin and Soft Tissue | 13 (10.2) |
| ▪ Gynecological | 9 (7.0) |
| ▪ Musculoskeletal | 8 (6.2) |
| ▪ Right Upper Quadrant | 7 (5.5) |
| ▪ Focused Assessment for Free Fluid (FAFF) | 3 (2.3) |
| ▪ Scrotal/Testicular | 3 (2.3) |
| ▪ Ocular | 2 (1.6) |
| ▪ Bowel/GI | 1 (0.8) |

**Table 2.** Utility of Sonography.

| **Table 2. Utility of Sonography** |  |
| --- | --- |
| **Variables** | **n=128** |
| **Provider Specialty, n (%)** |  |
| ▪ Internal Medicine | 76 (59.4) |
| ▪ Emergency Medicine | 52 (40.6) |
| **Sonographic Findings, n (%)** |  |
| ▪ Unremarkable Study | 53 (42.2) |
| ▪ Significant Finding of Clinical/Diagnostic Value | 62 (48.4) |
| ▪ Characterization of Clinical Status/Follow up | 12 (9.4) |
| **Outcome of Sonographic Assessment, n (%)** |  |
| ▪ Confirmed Working Diagnosis (Helped in Clinical Decision for Therapeutic Intervention) | 21 (16.4) |
| ▪ Referral to Higher Level of Care | 35 (27.3) |
| ▪ Ruled Out Main Differential Diagnosis | 9 (7.0) |
| ▪ Unremarkable Study of No Significant Clinical Value | 63 (49.3) |

Twelve distinct types of sonographic studies were performed with lung ultrasound being the most common (28.2%), followed by perinatal ultrasound (suspected or first trimester pregnancy (3.1%); and second and third trimester pregnancy (10.2%)). Renal, Cardiac, and skin and soft tissue studies constituted 10-12% each. Less utilized studies are shown in **Table 1**.

Regarding the findings of POCUS studies, approximately 1 in 2 studies (48.4%) revealed significant findings that helped to confirm a working diagnosis and subsequently formulate or optimize plan of management. 42.2% of studies were unremarkable and revealed no clinically relevant finding. It is important to note that unremarkable studies were often of clinical significance in helping to exclude various diagnoses. POCUS was utilized for follow up disposition in 9.4% of studies, mostly for pregnancy.

The impact of POCUS on clinical judgement and outcome was noted. Analysis of these outcomes revealed that therapeutic intervention was made based on US findings in 16.4% of encounters, while a referral to a higher level of care was recommended in 27.3% of cases. Moreover, reassurance with ruling out a main differential diagnosis was the case in 7% of encounters, whereas no specific intervention or recommendation was made in 49.2% of cases. An example of clinical impact following a POCUS study included prescribing antibiotics for POCUS proven infection. Referrals to higher level of care included referral to surgical services, antenatal care, and further medical assessment of suspicious mass.

## DISCUSSION

Our study found that POCUS can be used while delivering healthcare in a remote out-of-hospital resource-limited setting. POCUS was utilized by both IM and EM providers in pediatric and adult patients in this study. Utilization of POCUS by providers didn’t seem to impact their ability to effectively participate in treating large number of patients or impede patient flow in any meaningful way. In a rural resource-limited setting, it is essential to recognize that care is often delivered out of hospital in non-traditional settings such as schools. In these settings, basic services such as electricity are often unavailable. Hand-held POCUS devices are a reliable option as they utilize rechargeable bateries and portable smart phones.

As an image modality that can provide objective data, POCUS was helpful in identifying disease processes and guided therapies such as initiation of antibiotics for pneumonia and cellulitis. Additionally, providers used POCUS as part of their evaluation and decision-making process to refer patients to a higher level of care. Given that most patients in this setting were of low socio-economic status, the decision to refer patients to a higher level of care was challenging due to the financial burden of such referrals. Having objective evidence to guide this process helped both providers and patients by eliminating unnecessary transfers and giving the clinician data to help educate the patient about the reason for the referral, its urgency, and the services they needed to seek.

We found that POCUS can be of value while taking care of pregnant women. Approximately 10% of the POCUS studies helped guide future follow-up for women. This is particularly important due to the lack of established routine perinatal care and obstetric imaging in the area.

There is limited data published in the literature regarding utility of PCOUS in global health. In a systemic review, Baloescu et al^5^ noted that only 4.6% of academic research on POCUS focuses on the utility of POCUS in LMICs. Additionally, authors found no randomized control trials on this topic. Evaluated published studies were not of high quality; therefore, evaluation of the effect of POCUS implementation on morbidity or mortality could not be carried out. The review, however, described usability and diagnostic value of POCUS. Interestingly, only 3 of the 20 studies included in the review looked at utility of POCUS in an out-of-hospital or established clinic setting similar to our study.

Studies focusing on remote out-of-hospital or established clinic settings in LMICs are few. One study assessed telemedicine guided ultrasounds in rural Nicaragua^10^, whereas some other studies assessed ultrasound scans performed by trained physicians in Nicaragua and Haiti^11,12^, with all studies signaling that POCUS can positively influence medical decision and/or leading to beter clinical outcome. Additionally, to the best of our knowledge, no prior study looked at utilization of POCUS in an out-of-hospital resource-limited setting in Africa.

Few reports in the literature address the utility of POCUS in Africa. Most do so in a hospital setting and as part of medical education^13,14,15^. In a relatively large study assessing training curriculum of global health medical residents in POCUS in Malawi and Guyana, POCUS was reportedly useful in guiding diagnosis and management^15^. A study in a tertiary center in Cape Cost, Ghana concluded that non-cardiologists can be trained on cardiac POCUS successfully^14^. Similar results were obtained on another study in Kumasi, Ghana that showed that emergency medicine residents were successfully trained on POCUS^13^. These results are promising to consider the larger context and possible use of POCUS outside academic settings.

In general, there remains a lack of consensus guidance on best clinical practices when it comes to short-term medical missions especially in remote rural areas in LMICs^16^. When it comes to utilizing POCUS in those settings, more large-scale research is needed to assess the cost-effectiveness of implementing POCUS as part of the medical care delivered to this cohort.

## Limitations

Our study has few notable limitations. The retrospective design of the study limited amount and type of data that included in this analysis. More information about clinical characteristics of patients as well as follow up could improve the quality of US utility. Additionally, we recognize that while we were able to assess the utility of POCUS in out-of-hospital resource-limited settings, we were not able to assess for the impact of POCUS on long-term outcomes. An example of measurable impact is to collect feedback from patients about their understanding of their clinical conditions/diseases before and after sonographic imaging and discussion of results. Moreover, impact could also be assessed with follow up information after PCOUS including outcome of treatment or referral to higher level of care. These would be useful variables to be included in the future studies.

## CONCLUSIONS

POCUS can be utilized effectively in out-of-hospital resource-limited settings in rural Ghana with significant impact on clinical decision-making process. A wide range of scan types were be performed for various clinical indications by emergency medicine and internal medicine providers for adult and pediatric patients.

## Data Availability

All data produced in the present study are available upon reasonable request to the authors

